# A Randomized Controlled Trial Comparing Erector Spinae Plane and Transversus Abdominis Plane Blocks for Postoperative Analgesia after Elective Caesarean Section

**DOI:** 10.1101/2023.10.27.23297508

**Authors:** Aidan Eksteen, Janine Wagner, Thomas Kleyenstuber, Peter Kamerman

## Abstract

**Background:** Peripheral abdominal nerve blocks contribute to multimodal postoperative analgesia that enhances early recovery after caesarean sections. The transversus abdominis plane (TAP) block is an established technique that offers somatic abdominal analgesia. The erector spinae plane (ESP) block is a novel fascial plane block that may offer additional visceral analgesic effects. This study aimed to compare the postoperative analgesic efficacy of bilateral ultrasound-guided ESP blocks to TAP blocks in women undergoing caesarean sections under spinal anaesthesia.

**Methods:** Sixty-six ASA grade 1-3 (≥18 years) patients undergoing elective caesarean section under spinal anaesthesia were randomly allocated to receive either ESP blocks at the T9 vertebral level (*n* =33) or TAP blocks (*n* =33). The primary outcome measure was 24-hour cumulative morphine consumption. The secondary outcomes included the time taken to perform blocks, numeric rating scale (NRS) pain scores at 6- and 24-hours, effect of pain on activities of daily living (ADLs) and care for the newborn, time to first analgesic request, severity of opioid-related side effects, and patient satisfaction.

**Results:** There was no statistically significant difference in mean (SD) 24-hour cumulative morphine consumption between the ESP blocks and TAP blocks: 27 mg (14) vs 32 mg (15) (p=0.185). ESP blocks took longer to perform: 10.7 minutes (2.2) vs 9.0 minutes (2.5) (p<0.01). There were no significant differences in the other secondary outcomes.

**Conclusion:** ESP blocks did not significantly reduce postoperative analgesic requirements compared to TAP blocks after caesarean section under spinal anaesthesia. The ESP block did not demonstrate significant additional visceral analgesic effects.

**Trial Registration:** South African National Clinical Trial Registry (DOH-27-102022-5278), Pan African Clinical Trials Registry (PACTR202301645957324)

---

**What is already known on this topic:**

TAP blocks provide opioid-sparing somatic abdominal analgesia after caesarean sections. Emerging evidence suggests that ESP blocks may offer additional visceral analgesia for abdominal surgery, but limited data supports this for caesarean sections.

**What this study adds:**

Whilst ESP blocks may be used as a postoperative analgesic adjunct for caesarean sections, they not significantly reduce postoperative analgesic requirements or demonstrate clinically significant additional visceral analgesic effects compared to TAP blocks.

**How this study might affect research, practice or policy:**

This study suggests the degree of visceral analgesia offered by ESP blocks remains uncertain and further research in other patient populations is required to reach a consensus on the most superior peripheral abdominal nerve block technique for caesarean sections.

## Introduction

Caesarean sections are associated with moderate to severe pain in the postoperative period.^1^ Pain is a leading concern for patients undergoing caesarean sections and a major determinant of overall patient satisfaction.^2^ Pain is associated with numerous adverse neuroendocrine effects that lead to several postoperative complications. It can reduce the ability of the mother to care for the infant and can lead to chronic pain, opioid abuse and dependence.^3,4^ The caesarean section carries a relatively high incidence of poorly controlled postoperative pain globally.^3,5^

Obtaining maximal pain reduction with minimal side effects to the mother and infant can be achieved with a multimodal approach that includes peripheral abdominal nerve blocks as postoperative analgesic adjuncts.^4^ These blocks improve analgesia, are opioid-sparing, and can enhance early recovery after surgery.^6^ They are particularly beneficial when intrathecal morphine is omitted from a neuraxial technique, when other analgesic agents are contraindicated, and when general anaesthesia is used. Additionally, they may benefit patients at high risk for severe pain or chronic pain and may be used as a postoperative rescue analgesic technique.^6–8^

The transversus abdominis plane (TAP) block, described by Rafi in 2001, involves deposition of local anaesthetic between the transversus abdominis and the internal oblique muscles of the abdominal wall.^9^ It provides somatic analgesia of the abdominal wall but it lacks visceral analgesic effects.^10^ A meta-analysis by Champaneria *et al* ^11^ showed that TAP blocks reduce pain scores and opioid consumption thus several international guidelines recommend them as analgesic adjuncts after caesarean sections, especially when intrathecal morphine is omitted from a neuraxial technique.^4^

The erector spinae plane (ESP) block is a new fascial plane technique described by Forero in 2016, as a paravertebral technique that confers cranio-caudal spread of local anaesthetic from underneath the ESP muscles to the paravertebral space.^12^ It offers somatic and potentially visceral analgesia.^12,13^ It has gained popularity due to its simplicity, safety, and ability to provide segmental analgesia for a variety of surgical procedures. Originally shown to have analgesic benefits for procedures of the thorax, there is now emerging evidence that low thoracic and lumbar ESP blocks have analgesic and opioid-sparing effects for abdominal surgery.^8,12,14,15^ Its application in caesarean section analgesia has recently been studied with limited evidence suggesting it may be superior to the TAP block.^16,17^

There is a paucity of data on these regional techniques in low/middle-income countries (LMICs) such as South Africa. In addition, there is poor adherence to international guidelines that recommend using peripheral abdominal nerve blocks as analgesic adjuncts when intrathecal morphine is not used.^4^ To this end, this study hoped to improve analgesic practice by testing the hypothesis that the ESP block has additional visceral analgesic effects and offers superior analgesic efficacy to the TAP block. This study aimed to compare the postoperative analgesic efficacy of ESP blocks to TAP blocks in women undergoing caesarean section under spinal anaesthesia.

## Methods

### Ethical Approval

Ethical approval to conduct this study was obtained from the Human Research Ethics Committee of the University of the Witwatersrand, Johannesburg, South Africa on August 23, 2022 (Clearance number M220512). The study was registered before commencement with the South African National Clinical Trials Register on October 27, 2022 (DOH-27-102022-5278), URL: https://sanctr.samrc.ac.za/TrialDisplay.aspx?TrialID=8100 and Pan African Clinical Trials Registry on January 09, 2023 (PACTR202301645957324), URL: https://pactr.samrc.ac.za/TrialDisplay.aspx?TrialID=24267. Participant enrolment began on January 10, 2023. This study was conducted in accordance with the Declaration of Helsinki.^18^ Written informed consent was obtained from all participants.

### Study design and participants

This study was a prospective, single-centre, randomized controlled trial with blinding at the level of outcome assessors. The study population included sixty-six ASA I-III obstetric patients (≥18 years) who were scheduled for elective caesarean section to be performed under spinal anaesthesia at the Rahima Moosa Mother and Child Hospital (RMMCH) in Johannesburg, South Africa. A consecutive convenience sampling technique was used, and the study took place from January until March 2023 until sample realization was achieved.

Exclusion criteria included patients that had a: BMI ≥ 35 kg/m^2^; pre-existing chronic pain syndrome; contraindication to spinal anaesthesia, local anaesthetic or opioids; physical or mental impairment which would prevent them from being able to use a patient-controlled analgesia (PCA) pump postoperatively; spinal anaesthetic that was converted to general anaesthesia.

### Randomisation and blinding

Simple randomization was done with an allocation ratio of 1:1 using an online random number generator (GraphPad®, Dotmatics). Sealed, numbered, opaque envelopes were used sequentially to allocate patients. The sequence was generated by an independent assistant and envelopes were opened by the principal investigator (AE) once the patient was in the theatre. The outcome assessors collecting data for the primary and secondary outcomes remained blinded to the allocations throughout the study.

### Block procedure

After informed consent was obtained for the procedure, an intravenous catheter was placed and a pulse oximeter, non-invasive blood pressure monitor (NIBP), and electrocardiogram (ECG) were applied to the patient as recommended by the American Society of Anaesthesia (ASA). The spinal anaesthetic was conducted by an independent anaesthetic practitioner. An aseptic technique was used to perform spinal anaesthesia in the sitting position with a 26-Gauge pencil-point spinal needle (UlifeSA, Durban, RSA). After confirmation of clear cerebrospinal fluid, 9 mg of 0.5% hyperbaric bupivacaine with 10 mcg of fentanyl was introduced intrathecally. Patients were positioned supine with 15 degrees left lateral tilt and vital signs were observed at 1-minute intervals for the first 10 minutes and 2.5-minute intervals thereafter. An adequate spinal level was confirmed using an ice-cold pack. Surgery proceeded and after delivery of the foetus, a bolus of 2.5 IU of oxytocin was administered, followed by an infusion of 40 IU oxytocin in 1L Ringers Lactate at 125ml/hr.

At the end of surgery, either an ESP or TAP block was performed in theatre whilst the patient was being monitored as per ASA standards. All the ESP and TAP blocks were performed by AE to ensure standardization of technique. The ESP blocks were performed with patients in the right lateral decubitus position.

A curvilinear (2-5 MHz) ultrasound transducer (Mindray, Shenzhen, China) was placed longitudinally 2-3 cm lateral to the midline at the level of the ninth thoracic (T9) transverse process (TP). After sterilization of the skin, a 21-Gauge 110 mm echogenic block needle (SonoTap, Pajunk, Geisingen, Germany) was introduced in a cranio-caudal direction with an in-plane ultrasound approach. The needle was directed toward the TP and once on the TP, hydro-dissection with sterile saline was used to confirm adequate spread of fluid between the TP and erector spinae muscle group. After aspiration to exclude intravascular injection, 20 mLs of 0.25% bupivacaine with adrenaline was injected under vision. This procedure was then repeated on the other side of the spine.

The TAP blocks were performed in the supine position, using a curvilinear (2-5 MHz) ultrasound transducer (Mindray, Shenzhen, China). The posterior approach was used with the probe placed midway between the subcostal margin and the iliac crest in the anterior axillary line. The fascial plane between the internal oblique muscle and the transversus abdominis muscle of the abdominal wall was visualised until it tapered toward the quadratus lumborum muscle. After sterilization of the skin, a 21-Gauge 110 mm echogenic needle (SonoTap, Pajunk, Geisingen, Germany) was introduced from anterior to posterior, and confirmation of correct needle placement was made by separation of the muscle layers with sterile saline hydro-dissection. After aspiration to exclude intravascular injection, 20 mLs of 0.25% bupivacaine with adrenaline was injected under vision. This procedure was then repeated on the other side of the abdomen.

Patients were transferred from the theatre to the recovery room where their vital signs were monitored and acute complications from the blocks were excluded. AE prescribed a standardized postoperative analgesic script consisting of paracetamol 1g 6-hourly, ibuprofen 400mg 8-hourly, and a rescue anti-emetic of metoclopramide 10 mg 8-hourly.

In the recovery room, all patients received a morphine patient-controlled analgesia (PCA) pump (CADD Solis®, Smiths Medical ASD, Inc, USA). The PCA pump, with morphine in a concentration of 1mg/mL was set to allow a bolus dose of 1mL (1mg) with a 6-minute lockout and a maximum of 6 doses per hour. After discharge from the recovery room, patients were followed up by the blinded outcome assessors (members of the acute pain service at RMMCH) at 6-hours and 24-hours post block.

### Outcome measures

The outcome assessors obtained cumulative 24-hour morphine consumption and time to first analgesic request, from the PCA pump electronic record. Numeric rating scale (NRS) pain scores were obtained using an 11-point scale (0 no pain, 10 worst pain) at rest and movement (cough), at time intervals 0-hours, 6-hours, and 24-hours after block completion. Outcome assessors conducted interviews at 24-hours where they assessed patients for opioid-related side effects (nausea, vomiting, drowsiness, itching), the effect of pain on activities of daily living (ADLs) and care of the newborn, and patient satisfaction. The data was captured on a data collection form by the outcome assessors and transferred onto an Excel™ spreadsheet (Microsoft Corp, Redmond, WA, USA) by AE for analysis.

### Sample size calculation

The sample size was calculated in consultation with a biostatistician. Using the results from the study by Kamel *et al*, ^19^ which used the same interventions as in the present study, a pooled standard deviation of 1.83, and a difference in means of 1.29 mg morphine use was calculated. These data were then used to calculate a Cohen’s d value of 0.703, a large effect size. Assuming a power of 80%, a 5% significance level, and an effect size of 0.703, a minimum sample size of 33 participants per group was calculated.

### Statistical analysis

Data was analysed using R v4.2.2 (R Core Team, 2022). Continuous numeric data such as age, weight, dose of morphine, duration of surgery and anaesthesia were summarised as mean (standard deviation, SD), while median (interquartile range, IQR) was used for scores (numerical rating scales) and discrete data (doses of drugs administered). Nominal categorical data were summarised using counts (percentage, %). For all analyses, a two-sided hypothesis was assessed, with statistical significance taken at p < 0.05. The primary outcome was analysed according to a pre-specified plan, with a Welch’s t-test being used to compare 24-hour cumulative dose of morphine between the two groups. Consolidated Standards of Reporting Trials (CONSORT) guidelines were utilised in the study design, conduct of study, and presentation in line with EQUATOR network recommendations.^20^

## Results

### Study participants

Participant selection and enrolment are summarised in the CONSORT flow diagram (Figure 1). Two participants in the ESP group could not tolerate the intravenous catheter attached to their PCA device and requested early removal of the PCA device. Thus, 33 participants per group were analysed for all primary and secondary outcomes. There were no statistically significant differences in demographic, anaesthetic, and surgical characteristics of the two intervention groups (Table 1). No complications were observed with either block.

**Figure 1.**
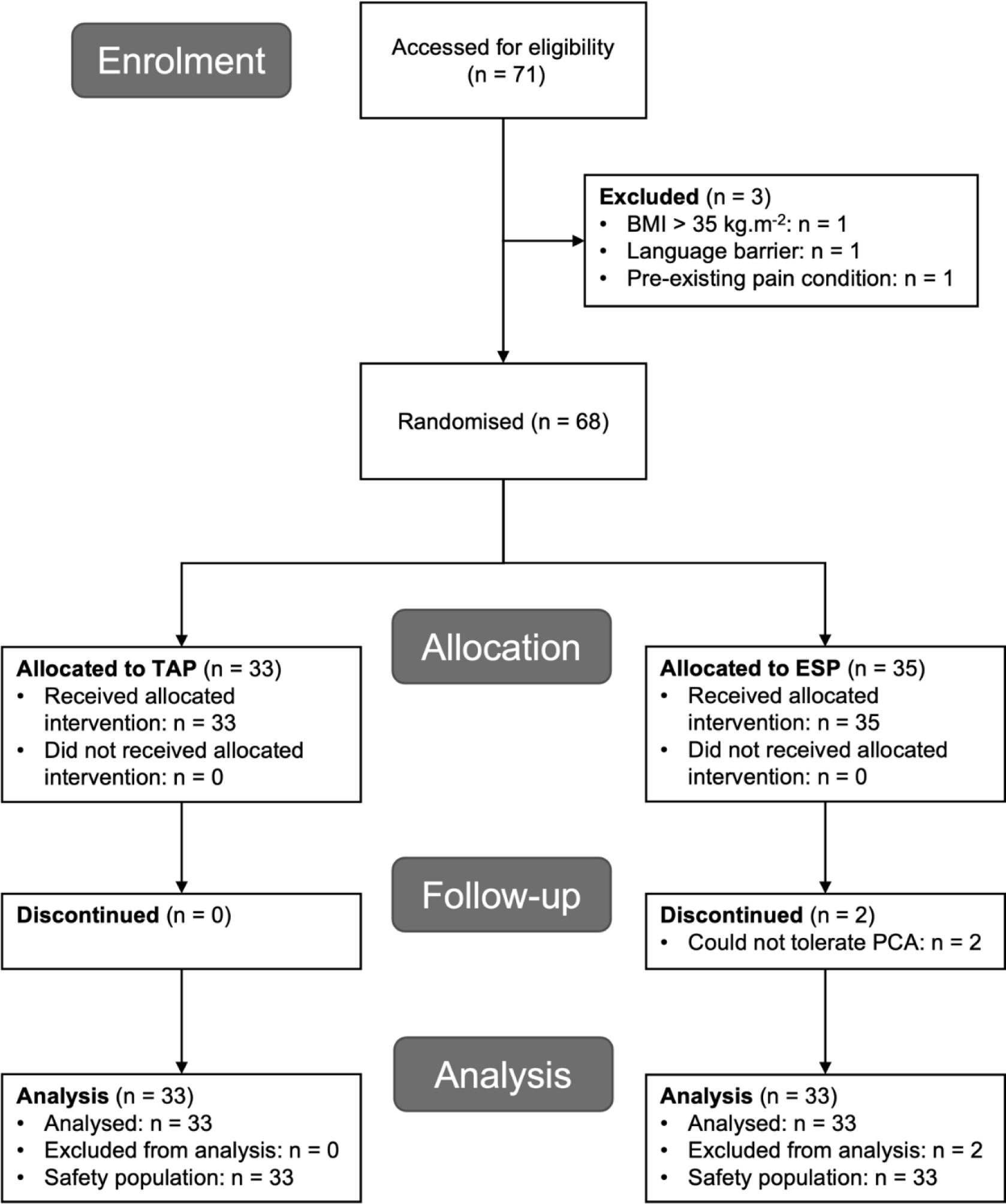
Consolidated Standards of Reporting Trials flow diagram of study participants. TAP: transversus abdominis plane block, ESP: erector spinae plane block.

**Table 1.** Patient, anaesthetic, and surgical characteristics.

|  | TAP | ESP |
| --- | --- | --- |
| Sample size, n | 33 | 33 |
| Patient characteristics |  |  |
| Mean age (SD) in years | 30.5 (5.0) | 31.1 (6.1) |
| Mean body mass index in kg/m<sup>2</sup> | 30.1 (3.2) | 28.5 (4.5) |
| Parity, n (%) |  |  |
| Multiparity | 31 (94) | 31 (94) |
| Primiparity | 2 (6) | 2 (6) |
| Previous abdominopelvic surgery, n (%) | 27 (82) | 30 (91) |
| Surgical and anaesthetic characteristics |  |  |
| Mean anaesthesia time (SD) in minutes | 89 (21) | 93 (17) |
| Mean regional block time (SD) in minutes | 9.0 (2.5) | 10.7 (2.2) |
| Mean surgical time (SD) in minutes | 45 (19) | 45 (18) |
| Type of skin incision, n (%) |  |  |
| Midline | 1 (3.0) | 3 (9.0) |
| Pfannenstiel | 32 (97) | 30 (91) |
| Adhesiolysis performed, n (%) | 12 (36) | 9 (27) |
| Parietal peritoneum closed, n (%) | 22 (67) | 17 (52) |
| Regional block complications |  |  |
| None, n (%) | 33 (100%) | 33 (100%) |
| Abbreviations: SD, Standard deviation; TAP, transversus abdominis plane block; ESP, erector spinae plane block |  |  |

### Primary outcome

The mean (SD) 24-hour cumulative dose of morphine was 32 (15) mg in the TAP group and 27 (14) mg in the ESP group (Figure 2), with a mean difference of 4.8 mg (95% CI: −2.3 to 11.9). The difference between the two interventions was not statistically significant (t-value = 1.34, degrees of freedom = 63.79, p= 0.185).

**Figure 2.**
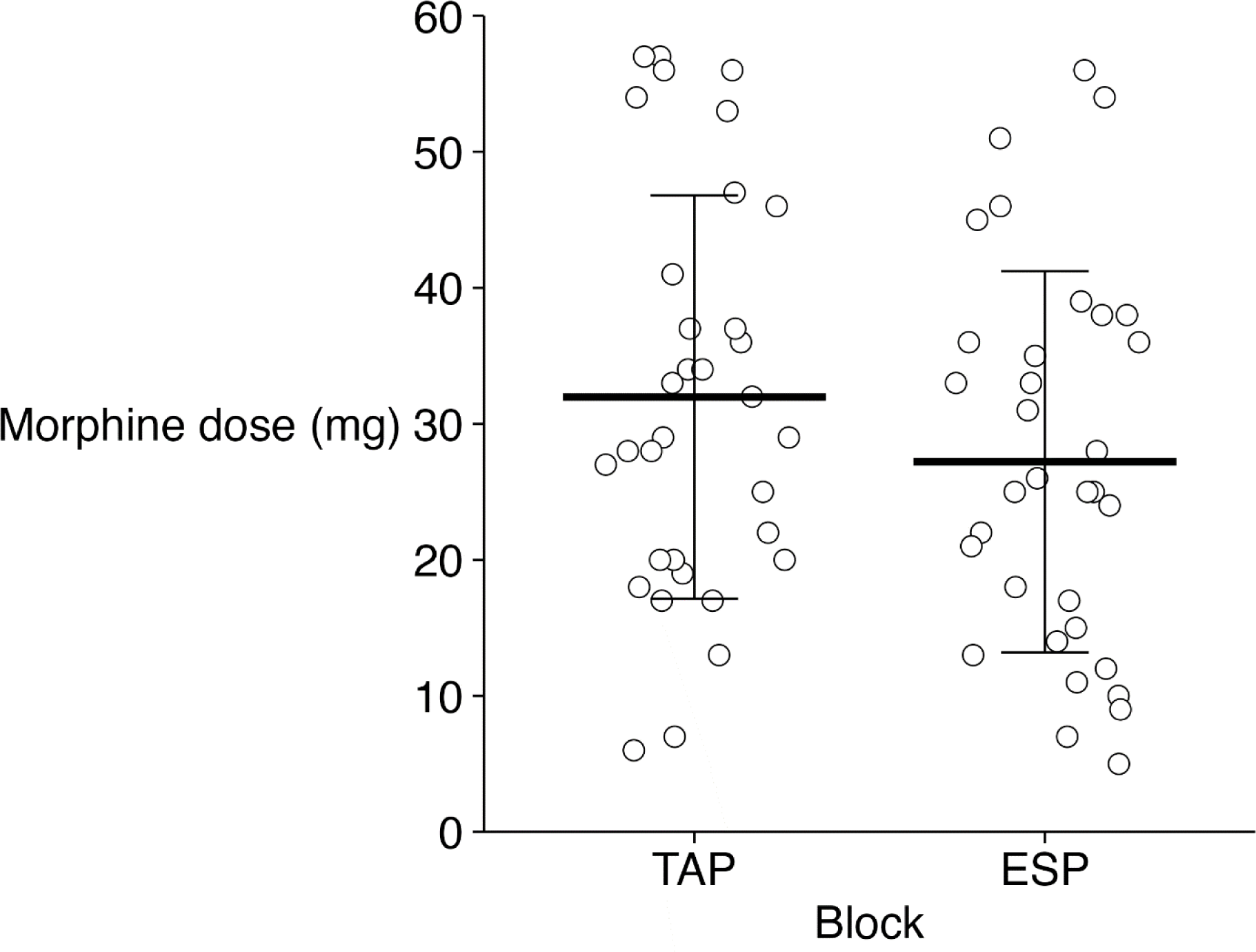
Mean (SD) 24-hour cumulative morphine dose administered via PCA device after caesarean section. TAP: transversus abdominis plane block, ESP: erector spinae plane block.

### Secondary outcomes

No statistically significant difference was found between the TAP and ESP groups for NRS pain scores at rest or with movement at 0, 6-, and 24-hours after block completion (Table 2). Additionally, there were no differences in the extent to which pain interfered with ADLs and care of the newborn (Table 2). Overall satisfaction with the analgesia provided was equally high in the groups (Table 2). There were no significant differences in the frequency or intensity of opioid-related side effects between the two groups (Table 3). There was no significant difference in median time (95 % CI) to first morphine PCA administration between the TAP group and the ESP group: 72 minutes (58 - 108) vs 84 minutes (65 - 122) (p=0.771). The mean (SD) time taken to perform the ESP blocks was marginally longer than the TAP blocks: 10.7 minutes (2.2) vs 9.0 minutes (2.5) (p<0.01).

**Table 2.** Pain scores, pain interference and patient satisfaction.

|  | TAP | ESP | p-value<sup>a,b</sup> |
| --- | --- | --- | --- |
| Sample size, n | 33 | 33 |  |
| Median NRS pain score (IQR) at time 0-hour<sup>c</sup> |  |  |  |
| At rest | 0 (0-0) | 0 (0-0) | 0.5 |
| With movement (cough) | 0 (0-0) | 0 (0-1) | 0.5 |
| Median NRS pain score (IQR) at time 6-hour<sup>c</sup> |  |  |  |
| At rest | 5 (3-8) | 6 (5-6) | >0.9 |
| With movement (cough) | 6 (5-8) | 7 (5-8) | 0.6 |
| Median NRS pain score (IQR) at time 24-hour<sup>c</sup> |  |  |  |
| At rest | 3 (2-4) | 2 (1-4) | 0.5 |
| With movement (cough) | 4 (2-7) | 3 (2-5) | 0.2 |
| Median pain interference score (IQR) with ADL<sup>d</sup> |  |  |  |
| Ability to move in bed | 4 (2-5) | 5 (3-7) | 0.14 |
| Ability to sleep | 3 (1-4) | 2 (0-4) | 0.5 |
| Ability to walk and stand | 3 (2-6) | 3 (1-5) | 0.14 |
| Ability to breastfeed | 1 (0-3) | 0 (0-3) | >0.9 |
| Ability to care for infant | 3 (1-5) | 3 (0-4) | 0.13 |
| Median satisfaction score (IQR) at 24-hour<sup>e</sup> | 9 (8-10) | 9 (8-10) | 0.8 |

Abbreviations: TAP, transversus abdominis plane block; ESP, erector spinae plane block; IQR, Interquartile range; NRS, Numeric rating scale; ADL, Activities of Daily living
a: p-value compares TAP to ESP
b: Wilcoxon sum rank test to compare NRS pain scores
c: 11-point numeric rating scale anchored at 0 (none) and 10 (severe)
d: 11-point numerical rating scale anchored at 0 (not at all) and 10 (extremely)
e: 11-point numerical rating scale anchored at 0 (not at all satisfied) and 10 (extremely satisfied)

**Table 3.** Opioid-related side effects.

|  | TAP | ESP | p-value<sup>a,b,c</sup> |
| --- | --- | --- | --- |
| Sample size, n | 33 | 33 |  |
| Nausea |  |  |  |
| Incidence, n (%) | 15 (45) | 11 (33) | 0.3 |
| Median (IQR) severity score<sup>d</sup> | 5 (2-6) | 2 (1-4) | 0.069 |
| Vomiting |  |  |  |
| Incidence, n (%) | 10 (30) | 10 (30) | >0.9 |
| Median (IQR) severity score<sup>d</sup> | 3 (1-8) | 2 (2-4) | 0.5 |
| Drowsiness |  |  |  |
| Incidence, n (%) | 24 (73) | 25 (76) | 0.8 |
| Median (IQR) severity score<sup>d</sup> | 4 (2-6) | 6 (3-7) | 0.2 |
| Itching |  |  |  |
| Incidence, n (%) | 11 (33) | 5 (15) | 0.085 |
| Median (IQR) severity score<sup>d</sup> | 5 (4-7) | 5 (3-8) | >0.9 |

Abbreviations: SD, Standard deviation; TAP, transversus abdominis plane block; ESP, erector spinae plane block
a: p-value compares TAP to ESP
b: Pearson's Chi-squared test to compare incidence
c: Wilcoxon sum rank test to compare median severity score
d: 11-point numeric rating scale anchored at 0 (none) and 10 (severe)

## Discussion

This study found that ESP blocks did not significantly reduce postoperative analgesic requirements compared to TAP blocks after elective caesarean section under spinal anaesthesia. There was no significant difference in the primary outcome of 24-hour cumulative morphine consumption. The time to first analgesic request; severity of postoperative pain; incidence and severity of opioid-related side effects; effect of pain on ADLs and care of the newborn; and patient satisfaction between the two blocks were comparable. The ESP blocks took longer to perform, likely due to the lateral positioning required. This is the first study to compare the postoperative analgesic efficacy of ESP to TAP blocks for caesarean sections, in a Southern African population. Importantly, patient-centred outcomes, such as opioid-related side effects, effects of pain on ADLs and patient satisfaction were evaluated for significance.

Peripheral abdominal nerve blocks remain important regional anaesthetic techniques for postoperative analgesia after caesarean sections as they have few complications and contribute to multimodal, opioid-sparing analgesia.^8^ The gold standard of spinal anaesthesia with intrathecal morphine has recently been challenged by a meta-analysis^1^ which found that a combination of ilioinguinal-iliohypogastric nerve blocks with TAP blocks, in patients receiving intrathecal morphine, was superior to those receiving intrathecal morphine alone. These techniques are under-utilised in LMICs especially where other analgesic recommendations, such as those made by the Procedure Specific Postoperative Pain Management (PROSPECT) working group, are not fully adhered to.^4,21^

There is limited data comparing these blocks after caesarean sections. Two other studies in publication have shown superior analgesic efficacy with the ESP block compared to the TAP block, however, this study’s findings are not consistent with those.^16,17^ Boules *et al* ^16^ found a 25mg lower median tramadol consumption with ESP blocks versus TAP blocks (p<0.01). They also found a mean visual analogue scale (VAS) pain score reduction of 0.32 in the ESP group versus the TAP group (p<0.01). Despite the difference in opioid consumption, there was no difference in patient satisfaction between the groups, which correlates with the findings of this study. Their study was different from this study on several grounds. Firstly, they utilised the lateral TAP approach as opposed to the posterior approach used in this study. The posterior approach has been shown to be more effective.^22^ Secondly, they did not assess whether there was a difference in opioid-related side effects given the difference in opioid consumption.

The study conducted by Malawat *et al* ^17^ also showed prolonged time to first analgesic request, lower total diclofenac consumption and lower VAS pain scores. In their study, a posterior TAP block was supplemented with rescue diclofenac boluses at a VAS score ≥ 40 mm, in contrast to this study, where patients could self-administer morphine for any quantity of pain. Additionally, they did not assess differences in patient satisfaction. Notably, a network meta-analysis by Wang *et al* ^23^ comparing six local anaesthetic techniques for caesarean section, did not find the ESP to be superior to the TAP block. They concluded that TAP blocks were overall the most effective local anaesthetic technique for postoperative analgesia.

The ESP block has been compared to the TAP block in several other abdominal procedures with variable outcomes. A study by Kamel *et al* ^19^ found lower morphine consumption in patients receiving ESP compared to TAP blocks after total abdominal hysterectomy, but there was no difference in postoperative nausea and vomiting. A study conducted by Warner *et al* ^24^ in patients presenting for laparoscopic hysterectomies, showed no significant difference in pain scores, opioid consumption, or patient satisfaction between ESP and TAP blocks.

A study by Hassanin *et al* ^25^ on emergency laparotomies, showed lower cumulative fentanyl consumption in the ESP vs TAP group but the study did not assess differences in opioid-related side effects or patient satisfaction. A meta-analysis by Liheng *et al* ^26^ comparing ESP and TAP blocks for abdominal surgery found ESP blocks lowered opioid consumption and pain scores statistically but did not reach clinically significant targets. Additionally, there were no differences in patient satisfaction.

There is conflicting evidence as to whether the ESP has additional visceral analgesic effects that make it superior to somatic blocks, such as the TAP block. The extent and reliability of ventral and sympathetic rami blockade in the paravertebral space, based on clinical, cadaveric and MRI imaging studies, is still unclear.^27^ While there is some evidence for potential visceral effects, there is a need to examine whether these statistically significant results equate to clinically significant outcomes. It could be argued that many of these results do not meet criteria for minimal clinically important differences (MCID) in pain scores and opioid consumption.^27,28^

A recent systematic review found the TAP block was equivalent to the Quadratus Lumborum (QL) block for pain scores and opioid consumption after caesarean section.^29^ Furthermore, Bakshi *et al* ^30^ showed ESP and QL blocks were equivalent for postoperative analgesia after caesarean sections. These studies suggest that none of these techniques are significantly superior to one another. Whilst more evidence is required to determine superiority between the two blocks, they can both be considered valuable tools in the regional nerve block armamentarium for caesarean section analgesia. This study emphasizes that formulation of a customised analgesic strategy with the selective use of peripheral abdominal nerve blocks is of utmost importance, regardless of the technique used.

Several limitations exist in our study. This single-centre trial utilised convenience sampling over a short period of time that could have introduced selection bias and the results may not be extrapolated to all patient populations. We were not able to test the dermatomal adequacy of the block with sensory testing due to the effects of the neuraxial anaesthesia at the time of the blocks. Due to staff rotations in the pain service, different outcome assessors conducted follow up on the patients in the study which may have introduced interviewer bias. Although several Quality of Recovery (QoR) outcomes were evaluated, further studies utilising standardised tests such as QoR-15 to assist in determining clinically significant outcomes are recommended. The concept of the minimum clinically important difference (MCID) for pain outcomes needs to be carefully explored for caesarean sections to determine adequate sample size calculation and relevance of results.

## Conclusion

This prospective, single-centre randomized controlled trial showed that bilateral, ultrasound-guided ESP blocks did not significantly reduce postoperative analgesic requirements compared to bilateral ultrasound-guided TAP blocks after elective caesarean section under spinal anaesthesia. The ESP block did not show significant additional visceral analgesic effects.

## Data Availability

All data produced in the present study are available upon reasonable request to the authors

## Acknowledgments

A special thanks to Candice Theunissen from Creatori Health (PTY) Ltd and Ronel De Haan from Gabler Medical (PTY) Ltd for assisting with a donation of consumables required for this study. A special thanks to the South African Society of Anaesthesiologists’ Jan Pretorius Research Fund for supplying financial assistance.

## Contributors

AE: conceptualised the study design, wrote the study protocol, collected data, and wrote the first draft of the article.

JW: provided research support and assisted with study design, data analysis, data presentation and article drafting.

TK: provided research support and assisted with study design, data analysis, data presentation and article drafting.

PK: provided research support and assisted with study design, data analysis, and data presentation and article drafting.

## References

1. Ryu C, Choi GJ, Jung YH, Baek CW, Cho CK, Kang H. Postoperative Analgesic Effectiveness of Peripheral Nerve Blocks in Cesarean Delivery: A Systematic Review and Network Meta-Analysis. J Pers Med. 2022;12(4). doi:10.3390/JPM12040634

2. Carvalho B, Cohen SE, Lipman SS, Fuller A, Mathusamy AD, Macario A. Patient preferences for anesthesia outcomes associated with cesarean delivery. Anesth Analg. 2005;101(4):1182–7. DOI 10.1213/01.ANE.0000167774.36833.99

3. Karlström A, Engström-Olofsson R, Norbergh KG, Sjöling M, Hildingsson I. Postoperative pain after cesarean birth affects breastfeeding and infant care. J Obstet Gynecol Neonatal Nurs. 2007;36(5):430–40. DOI 10.1111/J.1552-6909.2007.00160.X

4. Roofthooft E, Joshi GP, Rawal N, Van de Velde M, Joshi GP, Pogatzki-Zahn E, et al. PROSPECT guideline for elective caesarean section: updated systematic review and procedure-specific postoperative pain management recommendations. Anaesthesia. 2021;76(5):665–80. DOI 10.1111/ANAE.15339

5. Kintu A, Abdulla S, Lubikire A, Nabukenya MT, Igaga E, Bulamba F, et al. Postoperative pain after cesarean section: Assessment and management in a tertiary hospital in a low-income country. BMC Health Serv Res. 2019;19(1). DOI 10.1186/S12913-019-3911-X

6. Mitchell KD, Smith CT, Mechling C, Wessel CB, Orebaugh S, Lim G. A review of peripheral nerve blocks for cesarean delivery analgesia. Reg Anesth Pain Med. 2020;45(1):52–62. DOI 10.1136/RAPM-2019-100752

7. Bollag L, Lim G, Sultan P, Habib AS, Landau R, Zakowski M, et al. Society for Obstetric Anesthesia and Perinatology: Consensus Statement and Recommendations for Enhanced Recovery After Cesarean. Anesth Analg. 2021;132(5):1362–77. DOI 10.1213/ANE.0000000000005257

8. Chin KJ, McDonnell JG, Carvalho B, Sharkey A, Pawa A, Gadsden J. Essentials of Our Current Understanding: Abdominal Wall Blocks. Reg Anesth Pain Med. 2017;42(2):133–83. DOI 10.1097/AAP.0000000000000545

9. Rafi A. Abdominal field block: a new approach via the lumbar triangle. Anaesthesia. 2001;56(10):1024–6. DOI: 10.1111/J.1365-2044.2001.2279-40.X

10. Liu L, Xie YH, Zhang W, Chai XQ. Effect of Transversus Abdominis Plane Block on Postoperative Pain after Colorectal Surgery: A Meta-Analysis of Randomized Controlled Trials. Med Princ Pract. 2018;27(2):158–65. DOI 10.1159/000487323

11. Champaneria R, Shah L, Wilson MJ, Daniels JP. Clinical effectiveness of transversus abdominis plane (TAP) blocks for pain relief after caesarean section: a meta-analysis. Int J Obstet Anesth. 2016;28:45–60. DOI 10.1016/J.IJOA.2016.07.009

12. Chin KJ, El-Boghdadly K. Mechanisms of action of the erector spinae plane (ESP) block: a narrative review. Can J Anesth. 2021;68(3):387–408. DOI 10.1007/S12630-020-01875-2/FIGURES/3

13. Kwon HM, Kim DH, Jeong SM, Choi KT, Park S, Kwon HJ, et al. Does Erector Spinae Plane Block Have a Visceral Analgesic Effect?: A Randomized Controlled Trial. Sci Rep. 2020;10(1). DOI 10.1038/S41598-020-65172-0

14. Huang W, Wang W, Xie W, Chen Z, Liu Y. Erector spinae plane block for postoperative analgesia in breast and thoracic surgery: A systematic review and meta-analysis. J Clin Anesth. 2020;66. DOI 10.1016/J.JCLINANE.2020.109900

15. Viderman D, Aubakirova M, Abdildin YG. Erector Spinae Plane Block in Abdominal Surgery: A Meta-Analysis. Front Med. 2022;9:16. DOI 10.3389/FMED.2022.812531/BIBTEX

16. Boules ML, Goda AS, Abdelhady MA, Abu El SA, El-Azeem NA, Hamed MA. Comparison of analgesic effect between erector Spinae plane block and transversus abdominis plane block after elective cesarean section: A prospective randomized single-blind controlled study. J Pain Res. 2020;13:1073–80. DOI 10.2147/JPR.S253343

17. Malawat A, Verma K, Jethava D, Jethava D. Erector spinae plane block and transversus abdominis plane block for postoperative analgesia in cesarean section: A prospective randomized comparative study. J Anaesthesiol Clin Pharmacol. 2020;36(2):201–6. DOI 10.4103/JOACP.JOACP_116_19

18. World Medical Association. Declaration of Helsinki: Ethical Principles for Medical Research Involving Human Subjects. https://www.wma.net/policies-post/wma-declaration-of-helsinki-ethical-principles-for-medical-research-involving-human-subjects/ [Accessed 5th December 2021].

19. Kamel AAF, Amin OAI, Ibrahem MAM. Bilateral Ultrasound-Guided Erector Spinae Plane Block Versus Transversus Abdominis Plane Block on Postoperative Analgesia after Total Abdominal Hysterectomy. Pain Physician. 2020;23:375–82.

20. Schulz KF, Altman DG, Moher D, for the CONSORT Group. CONSORT 2010 Statement: updated guidelines for reporting parallel group randomised trials. https://www.equator-network.org/reporting-guidelines/consort/ [Accessed 5th December 2021].

21. Kahsay DT, Elsholz W, Habteselassie &, Bahta Z, Afr S, Analg JA, et al. Transversus abdominis plane block after Caesarean section in an area with limited resources. Medpharm. 2017;23(4):90–5. DOI 10.1080/22201181.2017.1349361

22. Faiz SHR, Alebouyeh MR, Derakhshan P, Imani F, Rahimzadeh P, Ashtiani MG. Comparison of ultrasound-guided posterior transversus abdominis plane block and lateral transversus abdominis plane block for postoperative pain management in patients undergoing cesarean section: a randomized double-blind clinical trial study. J Pain Res. 2018;11:5. DOI 10.2147/JPR.S146970

23. Wang J, Zhao G, Song G, Liu J. The Efficacy and Safety of Local Anesthetic Techniques for Postoperative Analgesia After Cesarean Section: A Bayesian Network Meta-Analysis of Randomized Controlled Trials. J Pain Res. 2021;14:1559. DOI 10.2147/JPR.S313972

24. Warner M, Yeap YL, Rigueiro G, Zhang P, Kasper K. Erector spinae plane block versus transversus abdominis plane block in laparoscopic hysterectomy. Pain Manag. 2022;12(8):907–16. DOI 10.2217/PMT-2022-0037

25. Hassanin AAM, Ali NS, Elshorbagy HM. Efficacy of ultrasound-guided transversus abdominis plane block versus erector spinae plane block for postoperative analgesia in patients undergoing emergency laparotomies: A randomized, double-blinded, controlled study. Egypt J Anaesth. 2022;38(1):521–8. DOI 10.1080/11101849.2022.2124660

26. Liheng L, Siyuan C, Zhen C, Changxue W. Erector Spinae Plane Block versus Transversus Abdominis Plane Block for Postoperative Analgesia in Abdominal Surgery: A Systematic Review and Meta-Analysis. J. Investig. Surg. 2022;35(9):1711–22. DOI 10.1080/08941939.2022.2098426

27. Hussain N, Brull R, Noble J, Weaver T, Essandoh M, McCartney CJL, et al. Statistically significant but clinically unimportant: A systematic review and meta-analysis of the analgesic benefits of erector spinae plane block following breast cancer surgery. Reg Anesth Pain Med. 2021;46(1):3–12. DOI 10.1136/RAPM-2020-101917

28. Myles PS, Myles DB, Galagher W, Boyd D, Chew C, MacDonald N, et al. Measuring acute postoperative pain using the visual analog scale: the minimal clinically important difference and patient acceptable symptom state. Br J Anaesth. 2017 Mar 1;118(3):424–9. DOI 10.1093/BJA/AEW466

29. El-Boghdadly K, Desai N, Halpern S, Blake L, Odor PM, Bampoe S, et al. Quadratus lumborum block vs. transversus abdominis plane block for caesarean delivery: a systematic review and network meta-analysis*. Anaesthesia. 2021;76(3):393–403. DOI 10.1111/ANAE.15160

30. Bakshi A, Srivastawa S, Jadon A, Mohsin K, Sinha N, Chakraborty S. Comparison of the analgesic efficacy of ultrasound - guided transmuscular quadratus lumborum block versus thoracic erector spinae block for postoperative analgesia in caesarean section parturients under spinal anaesthesia — A randomised study. Indian J Anaesth. 2022;66(4):S213–S219. DOI 10.4103/IJA.IJA

